# Dietary Nicotianamine as a Factor in International Variations of Mortality from Covid-19

**DOI:** 10.1101/2020.10.15.20213470

**Authors:** Charles E. Day

**Affiliations:** Audax, Leitchfield, KY 42754

## Abstract

Nicotianamine is a compound found only in plants that strongly binds to and potently inhibits at nanomolar concentrations the angiotensin converting enzyme 2 (ACE2) which is the human cell receptor for the coronavirus SARS-CoV-2. Because of its ACE2 binding activity it can potentially reduce mortality from Covid-19. This report explores the inverse association of dietary nicotianamine intake with mortality from Covid-19 from numerous countries. The data support a probable dramatic effect of nicotianamine in reducing coronavirus mortality. The best-case scenario of consuming a diet high in nicotianamine is a 100-fold reduction in Covid-19 mortality. Worst case is that one consumes a more healthful diet that is less expensive and more environmentally friendly.

## Introduction

By the first week of October 2020, over one million people worldwide had died from the Covid-19 pandemic in more than 200 countries with the United States being hit hardest and accounting for almost a quarter of the total global deaths. One remarkable aspect of the pandemic is the extreme variation of the Covid-19 mortality rates among different countries, ranging from over 1000 deaths per million population down to zero deaths per million.

Numerous factors such as lockdowns, social distancing, and mask mandates to failures to test and/or report data have been proffered to explain this marked intercountry variation in Covid-19 mortality rates. Although all may play a role, none can adequately explain all the observed variance. However, as first proposed in March of 2020, a little-known dietary factor can potentially explain a significant amount of the observed mortality variations (1). This dietary compound is nicotianamine.

Nicotianamine was first isolated from green tobacco leaves, *Nicotiana tabacum L*., in 1971 by Noma, Noguchi, and Tamaki (2). From 70kg of green leaves they isolated 209mg of a previously unknown, water-soluble amino acid which they named nicotianamine. Although nicotine also occurs in tobacco, this addictive substance bears no structural or functional relationship to nicotianamine. Additionally, this compound should not be confused with the vitamin B3 nicotinamide. They merely have look-a-like names but are otherwise structurally and functionally distinct.

Virtually all higher plants make nicotianamine since this material serves a vital function for them by chelating heavy metals like iron and zinc which can be more readily extracted and utilized from the soils the plant grows in. Therefore, most of the research conducted on nicotianamine has been in the plant sciences usually aimed toward improving the agricultural or agronomic characteristics of various crop plants. Plants synthesize nicotianamine by condensing three molecules of S-adenosyl-methionine via the enzyme nicotianamine synthase to make one molecule of nicotianamine. Much research has centered around transferring nicotianamine synthase genes to enhance synthesis of nicotianamine to improve plant qualities (3).

Although most plants make nicotianamine, each makes it in different amounts. Research on the isolation and quantitation of nicotianamine in vegetables and plant derived foods has been conducted almost exclusively by Japanese scientists. In three different reports (4-6), they have published quantitative amounts for nicotianamine in 166 plant foods. They were unable to detect any nicotianamine, zero, in only four of those foods which were asparagus, rice, wheat, and corn. Some of the foods with the highest content of nicotianamine were soybeans (45mg/100g), lentils (36mg/100g) and chickpeas (31.5mg/100g).

Nicotianamine potently binds to and inhibits human angiotensin converting enzyme 2 (ACE2) at nanomolar concentrations (7). The IC50 inhibitory concentration in vitro is 84 nM. The coronavirus SARS-CoV-2 avidly binds human ACE2 to gain entrance to human cells where it replicates profusely and causes the disease Covid-19. The possibility exists that dietary nicotianamine in sufficient quantities could compete with SARS-CoV-2 for binding to human ACE2 to ameliorate Covid-19. Beyond the fact that nicotianamine is soluble in water, absolutely nothing is known about human absorption, distribution, metabolism, excretion, and pharmacokinetics of this compound. However, it may be possible to obtain a crude estimate of how much dietary nicotianamine would be required to be minimally effective at ameliorating Covid-19.

If 5mg of nicotianamine were evenly distributed in a volume of water contained in a 70kg person, then the concentration would be 336 nM which would likely be sufficient to have a negative impact on SARS-CoV-2 binding to ACE2. As a working hypothesis, 5mg/day of dietary nicotianamine will be used as a minimal daily intake to favorably influence Covid-19.

Since nicotianamine is made only by plants, any animal derived food has zero amounts of this material. Also, wheat and corn contain no nicotianamine, and any foods derived from them also contain none. Eliminate all meats, seafoods, eggs, dairy products, and foods made from wheat and corn, from the American diet and Americans would quickly die from starvation. The American people consume, at best, very negligible amounts of nicotianamine in their low vegetable and fruit diet.

This report will examine Covid-19 mortality data from several different populations that consume more than 5mg/day of nicotianamine from different dietary sources and compare mortality data from countries which consume virtually none of this compound like the United States.

## Methods

All coronavirus mortality data are taken from the www.worldometers.info/coronavirus/ database which is updated daily. For this report, the database was last accessed on October 8, 2020. Five different country groups which consume as staples in their daily diets foods which contain high levels of nicotianamine, at least 5mg/day, are examined. The groups of countries evaluated are those that consume the highest levels of 1) soybeans, 2) lentils, 3) chickpeas, 4) buckwheat, and 5) barley.

## Results and Discussion

### No Nicotianamine Diets

Fruit, vegetable, and whole grain consumption is far below recommended levels in most affluent countries, and protein is largely from relatively expensive animal sources such as meat, seafood, eggs, and dairy products. Wheat and corn are used for popular food products like pastas, breads, chips, cookies, and cakes. All animal derived foods and wheat and corn contain zero nicotianamine. Pizza is one of the most popular foods in America and consists mostly of wheat flour, cheese, and meat, none of which contain any nicotianamine. Indeed, the usual American diet is virtually devoid of nicotianamine. Similarly, the diets of most western European nations contain little if any nicotianamine.

On October 8, 2020 the Covid-19 mortality for the United States was 654 deaths per million population. The rates in the western European nations of Great Briton, Spain, France, Italy, and Belgium were 625, 696, 497, 597, and 870, respectively, with an average mortality rate of 656 for these countries whose populace consumes a diet with very little to no nicotianamine.

### Soybeans

Of the foods for which nicotianamine content has been measured, soybeans contain one of the highest levels at 45mg/100g. The country consuming the most soybeans in the world is Japan. In 2014, on average, Japanese consumed 59.4g/day of soybeans which equates to a daily intake of 26.7mg nicotianamine. Japanese over 70 years ate 71.6g daily containing 32.2mg nicotianamine (8).

After Japan, in decreasing order, countries which ate the most soybeans in 2007 were South Korea, China, Nigeria, Vietnam, and Thailand (8). On October 8, 2020, starting with Japan, the cumulative Covid-19 mortalities in deaths per million population for those countries were 13, 8, 3, 5, 0.4, and 0.8, respectively, with the average for the six countries consuming the most soybeans being 5. This is lower than the mortality rate for Covid-19 in the United States by a factor of 130. This means that for every 130 Americans who die from Covid-19, only one person dies in countries eating the most soybeans.

American farmers produced 834 pounds of soybeans for every man, woman, and child in America in 2018 (9). Most of those which were not sold to soy consuming nations were crushed to extract their oil for consumption by Americans. The by-product after oil extraction is soybean meal which is used mostly for chicken and pig feeds. Since nicotianamine is water soluble, all of it remains in the soybean meal with none in the oil. Americans eat the oil and feed the meal to livestock.

Soybean meal production in the United States is 45.64 million metric tons annually (10) which equates to 382g per person per day, all of which is fed to livestock. Soybean meal for human consumption is sold as soy grits. Two ounces, 56.75g, of soy grits contains 25.5mg of nicotianamine. If purchased at the feed mill as soybean meal, this amount costs about a penny a day.

If nicotianamine really does reduce mortality from Covid-19, it is tragic that more than 200,000 American lives might have been saved by giving each of them a penny’s worth of soybean meal each day.

### Lentils

At 36mg/100g lentils also contain high amounts of nicotianamine. Lentils are also very high in protein and very cheap. Since adequate protein is necessary for human survival, and animal protein is relatively expensive, the poorest of nations are forced to depend on plant proteins, largely lentils, for their survival needs. The seven nations getting the highest percentage of their protein from pulses (lentils and peas) are the very poor nations Burundi, Rwanda, Uganda, Kenya, Comoros, Haiti, and Eritrea (11) which get 55, 38, 20, 20, 18, 18, and 18 percent, respectively, of their protein from pulses.

Lentils provide 25.8g protein per 100g (12). Assuming a daily protein requirement of 56g/day, it would require 119g of lentils to meet 55% of the daily protein needs of a Burundian which would provide 42.8mg/day nicotianamine. An Eritrean who gets 18% of his daily protein requirement from pulses consumes 14.1mg nicotianamine per day from lentils.

Total mortality rates through October 8, 2020 for the seven countries, which are Burundi, Rwanda, Uganda, Kenya, Comoros, Haiti, Eritrea, that consume the highest amounts of lentils, are 0.08, 2, 2, 14, 8, 20, and 0, respectively, averaging 6.6 deaths per million people.

### Chickpeas

The chickpea is highly consumed in many countries. Think hummus and falafels. In 2004 the top chickpea consuming nations were Turkey, India, Myanmar, Jordan, and Pakistan with Turkey consuming the most at 6.65kg/person/year (13). This amount (18.2g/day) provides 5.7mg/day nicotianamine. The total Covid-19 mortalities as of October 8, 2020 for these countries, respectively, were 102, 76, 9, 13, and 29 averaging 38.2 deaths per million.

In addition to chickpeas, these countries, especially India and Pakistan, consume significant quantities nicotianamine from lentils as well.

### Buckwheat

Although buckwheat does not contain nicotianamine per se, it does contain high amounts (30mg/100g) of a closely related analog, 2”-hydroxynicotianamine, which potently inhibits ACE

1 with an IC50 value of 80 nM (14). Presently, it has not been shown to inhibit ACE2. However, it is not unreasonable to make that assumption.

Countries consuming the most buckwheat are Russia, Ukraine, and Kazakhstan (15). Russians consume the most at 15kg/person/year. This amount equates to a daily intake of 12.3mg/day of hydroxynicotianamine.

Covid-19 mortalities for Russia, Ukraine, and Kazakhstan are 150, 105, and 93, respectively, with an average of 116 deaths per million people.

### Barley

Barley grain contains 6.7mg/100g nicotianamine which is soluble in water. Greatly simplified, beer can be broadly viewed as a water extract of barley. If 2.2 pounds of barley were used to make a gallon of beer, and if all the nicotianamine were extracted into the water, then the resulting beer would contain 17.7mg nicotianamine per liter.

The countries which consume the most beer in the world are Czechia, Austria, and Germany (16). Based on that consumption and assuming a 17.7mg/liter nicotianamine content, their daily per capita intake of nicotianamine from beer alone is 11.0, 6.4, 5.8 mg/day/adult for Czechia, Austria, and Germany, respectively. Their total Covid-19 mortalities through October 8, 2020 are 77, 92, and 115, respectively, averaging 94.7 deaths per million population.

### Overview

Several foods contain especially large quantities of nicotianamine or hydroxynicotianamine. These foods include soybeans, lentils, chickpeas, buckwheat, and barley. Invariably, when one examines the total mortalities from Covid-19 from the countries which consume the most of these foods, a dramatic reduction in mortality is observed versus those countries where very little nicotianamine is consumed (Table 1).

**Table 1.** Average Covid-19 Mortality Rates from Countries which Consume the Most of These Foods Containing High Levels of Nicotianamine (NA)

| **High NA Food** | **Total Countries** | **Average Covid-19 Mortality**<br>**(deaths/million)** | **Upper Level of NA**<br>**from Selected Food**<br>**(mg/day/person)** |
| --- | --- | --- | --- |
| **None** | **6** | **656** | **<1** |
| **Soybeans** | **6** | **5.0** | **32.2** |
| **Lentils** | **7** | **6.6** | **42.8** |
| **Chickpeas** | **5** | **38.2** | **5.7** |
| **Buckwheat** | **3** | **116** | **12.3** |
| **Barley (beer)** | **3** | **94.7** | **11.0** |

Many more countries probably could be added to the list of countries discussed above and summarized in Table 1. However, only countries for which quantitative consumption data are available for these foods are considered in this investigation. Many more Asian, Middle Eastern, and African nations likely consume diets high in nicotianamine in addition to those documented to consume such diets as those considered in this report.

## Conclusions

Nicotianamine is a heavy metal chelator, especially iron and zinc, made by plants that potently, at nanomolar concentrations, binds to and inhibits the ACE2 protein in human cells. Since ACE2 is the protein to which the coronavirus causing Covid-19 binds to gain entry to human cells, the possibility exists that nicotianamine may compete with the virus for binding to ACE2. If so, then high daily intake of dietary nicotianamine could possibly interfere with infectivity and/or level of coronavirus infections. It is not un-noticed that nicotianamine could be a zinc ionophore that additionally inhibits intracellular coronavirus replication.

To explore this possibility of potentially salutary benefits of this phytochemical against SARS-CoV-2, Covid-19 mortality rates in countries with documented consumption of foods high in nicotianamine were compared with those in countries that consume diets virtually devoid of nicotianamine. The average Covid-19 mortality rates in those countries consuming the most nicotianamine versus those eating the least are 100-fold lower (Table 1).

Dietary nicotianamine is one factor, among many, that can potentially explain the dramatic variations in Covid-19 mortality rates among dozens of countries. Indeed, if it possesses the activity against Covid-19 suggested by these data, then the need for vaccines and therapeutics may be obviated by a simple, safe and extremely inexpensive change in diet, and the fear of Covid-19 would disappear.

## Data Availability

Covid-19 mortality data available at worldometers.org/coronavirus/

## References

1. C.E. Day. 2020. How Not to Die from Covid-19. (www.booksbyday.com)

2. M. Noma, M. Noguchi, and E. Tamaki. 1971. A new amino acid, nicotianamine, from tobacco leaves. Tetrahedron Letters 22: 2017–2020.

3. T. Nozoye. 2018. The nicotianamine synthase gene is a useful candidate for improving the nutritional qualities and Fe-deficiency tolerance of various crops. Frontiers Plant Science Volume 9, Article 340. (www.ncbi.nlm.nih.gov/pmc/articles/PMC5881101)

4. H. Izawa, N. Yoshida, N. Shiragai, and Y. Aoyagi. 2008. Nicotianamine content in various beans and its inhibition activity of angiotensin-1 converting enzyme. Nippon Shokuhin Kagaku Kogaku Kaishi 55: 253–257. (www.jstage.jst.go.jp/article/nskkk/55/5/55_5_253/_pdf/-char/en)

5. H. Izawa and Y. Aoyagi. 2012. Nicotianamine contents among vegetables and their inhibitory activity of angiotensin-1 converting enzyme. Nippon Shokuhin Kagaku Kogaku Kaishi 59: 348–353. (www.jstage.jst.go.jp/article/nskkk/59/7/59_348/_pdf/-char/en)

6. H. Yamaguchi and R. Uchida. 2012. Determination of nicotianamine in soy sauce and other plant-based foods by LC-MS/MS. Journal Agric. Food Chem. 60: 10000–10006.

7. S. Takahashi, T. Yoshida, K. Yoshizawa-Kumagaye, and T. Sugiyama. 2015. Nicotianamine is a novel angiotensin-converting enzyme 2 inhibitor in soybean. Biomedical Research (Tokyo) 36: 219–224. (www.jstage.jst.go.jp/article/biomedres/36/3/36_219/_article)

8. www.otsuka.co.jp/en/nutraceutical/about/soylution/encyclopedia/consumption.html

9. https://en.wikipedia.org/List_of_countries_by_soybean_production

10. https://www.indexmundi.com/agriculture/?country=us&commodity=soybeanmeal&graph=production

11. http://legumelab.msu.edu/uploads/files/Maredia%20Presentation%20-%20Global%20Pulse%20Production%20and%20Consumption%20Trends.pdf

12. https://nutritiondata.self.com/facts/legumes-and-legume-products/4337/2

13. https://www.researchgate.net/publication/236667218_Uses_Consumption_and_Utilization%20(1).pdf

14. Y. Aoyagi. 2006. An angiotensin-1 converting enzyme inhibitor from buckwheat (Fagopyrum esculentum Moench) flour. Phytochemistry 67: 618–621.

15. http://en.wikipedia.org/wiki/Kasha

16. https://en.wikipedia.org/wiki/List_of_countries_by_beer_consumption_per_capita

